# Genomic profiling reveals molecular heterogeneity in patients with Richter transformation (RT) and chronic lymphocytic leukemia (CLL)

**DOI:** 10.1101/2025.03.15.25324025

**Authors:** Shulan Tian, Hanyin Wang, Sameer A. Parikh, Yuanhang Liu, Helen Jin-Lee, Erik Jessen, Eric W. Klee, Yucai Wang, Fan Leng, Min Shi, Cinthya Zepeda-Mendoza, Rong He, Saad J. Kenderian, Linda B. Baughn, Daniel L. Van Dyke, Paul J. Hampel, Neil E. Kay, Esteban Braggio, Susan L. Slager, Huihuang Yan, Wei Ding

**Author notes:** Correspondence (H.Y.); (W.D.). These authors contributed equally to this work (Shulan Tian, Hanyin Wang).

## Abstract

Richter transformation (RT) represents the development of an aggressive lymphoma in chronic lymphocytic leukemia (CLL). Patients with RT and relapsed CLL have poor outcomes. Yet, the extent of molecular differences between the two entities has not been fully explored. In this pilot study, we conducted RNA-seq and targeted panel sequencing of nodal tissues from 12 patients, including seven with RT and five with CLL. Analysis of RNA-seq data revealed two major clusters, with five RT in cluster C1 and the remaining two RT and all five CLL in C2. Within C2, one of the CLL ultimately developed RT; it showed more similarity to the two RT than to the other CLL in expression profile, suggesting the presence of expression signature for RT prior to the clinical diagnosis. In addition, differentially expressed genes, the majority of which showed higher expression in C1 relative to C2, were enriched in pathways known to be important for CLL pathogenesis or transformation. Deconvolution of the bulk RNA-seq data revealed major differences in cellular composition between the two clusters, notably tumor B cells, macrophages M1, and CD8+ T cells. Furthermore, by targeted sequencing, we identified 51 genes that carried recurrent copy number alterations (CNAs) preferentially occurring in either cluster. Over 80% of these CNAs occurred in C2, mainly gains of 17q12q25 in CLL. Patients in C1 had shorter overall survival (median 11 months) compared to those in C2 (median 36 months). Together, our findings highlight noticeable differences in transcriptomic and genomic alterations between CLL versus RT.

**Simple Summary:** Chronic lymphocytic leukemia (CLL) is a type of blood cancer, representing the most common leukemia in the Western countries. A small percent of patients with CLL develop into an aggressive lymphoma, most commonly diffuse large B-cell lymphoma (DLBCL), a process known as Richter transformation (RT). In RT, previous studies have identified specific somatic events and molecular subtypes. A deep understanding of the molecular events driving CLL to RT will accelerate the development of therapeutic strategies. Using RNA-seq and targeted sequencing, this study revealed heterogeneity of gene expression, copy number alterations, and dysregulated pathways in a cohort of 12 RT/CLL.

## 1. Introduction

Richter transformation (RT) represents the development of an aggressive lymphoma, most commonly diffuse large B-cell lymphoma (DLBCL), in chronic lymphocytic leukemia (CLL) (1, 2). Very often, RT represents true transformation from CLL B cells, which has a poor prognosis with a median overall survival of <12-24 months (3, 4). In contrast, clonally unrelated RT derived from a different clone has better prognosis (1, 5). Previously, we observed clinically durable response to pembrolizumab in patients with RT who experienced progression following ibrutinib treatment, but less so for patients with relapsed CLL (6). However, the factors contributing to the differential response remain largely unknown.

The molecular events driving CLL to RT are not fully understood (4, 7). Compared to CLL, RT overall has a higher mutational burden (7, 8), particularly copy number (CN) alterations (CNAs) (7). In RT, the most common genomic alterations include *TP53* inactivation mutations/deletions (as part of del(17p)) (8-11), deletion of the *CDKN2A*/*B* at 9p21 (8-11), as well as activation of *MYC* (8, 9) and *NOTCH1* (8, 9). These genes play key roles in tumor suppression, cell cycle control and cell proliferation. A few recent studies utilized longitudinal samples (7), paired circulating CLL and RT biopsies (8), or computational deconvolution of CLL and RT cells from patients with RT (4). These studies have identified additional ∼20 genes with somatic mutations and deletions in RT, such as *PTPRD* and *TRAF3* (8), *ARID4B*, *CDKN1A*/*1B* and *IRF4* (7), as well as *CCND3*, *DNMT3A* and *IRF2BP2* (4).

Fluorescence in situ hybridization (FISH) and DNA sequencing based studies have identified numerous CNAs in RT, frequently deletions of 17p (including *TP53*) (8, 10), deletions of 9p21 (including *CDKN2A/B*) (10), deletions of 13q14 (10), trisomy 12 (8, 10), and amplification of 8q (including *MYC*) (8, 12), where deletion of 9p21 rarely occurs in the CLL phase (4, 10). Finally, whole-genome duplication (4) and chromothripsis (4, 7) were identified exclusively in RT.

We hypothesized that early detection of genomic aberrations associated with a high risk of transformation is critical for developing potentially targeted therapeutic strategies (10, 13, 14). Here we performed RNA-seq and Tempus gene panel sequencing (see below in Methods) of nodal tissues from a cohort of patients with CLL (n=5) and RT (n=7). We identified two major clusters with distinct expression profiles, with cluster C1 including five of the seven RT cases and C2 including the other two RT and all five CLL cases. We also identified 51 genes with CNAs that preferentially occurred in either cluster, with the majority representing 17q gains in CLL within C2.

## 2. Material and Methods

### 2.1. CLL patients

Twelve patients with CLL who had lymph node biopsies for evaluation of clinical progression at Mayo Clinic were included in this study (Table S1). Based on pathological findings (Figure 1), seven patients had confirmed RT (large cell lymphoma histology, sample ID starting with RT). The other five had CLL (sample ID starting with CLL), of whom CLL244 developed RT 14 months after tissue biopsy showed CLL (Figure 1). For the treatment history, two patients (RT207 and CLL246) did not receive ibrutinib treatment before lymph node biopsy. Of the other ten who received ibrutinib treatment for a median of 23 months prior to the lymph node biopsy, one (RT256) was discontinued after 42 months of treatment and another one (RT037) was on venetoclax at the time of biopsy (Table S1). Chromosomal abnormalities, including del(11q), del(13q), del(17p) and trisomy 12, were based on fluorescence in situ hybridization (FISH) (15). Overall survival (OS) was defined as the time from initiation of ibrutinib treatment to death or the last follow-up (16). The study was approved by the Institutional Review Board at Mayo Clinic. Samples were collected as part of routine care of all patients and used for both targeted DNA and RNA sequencing.

**Figure 1.**
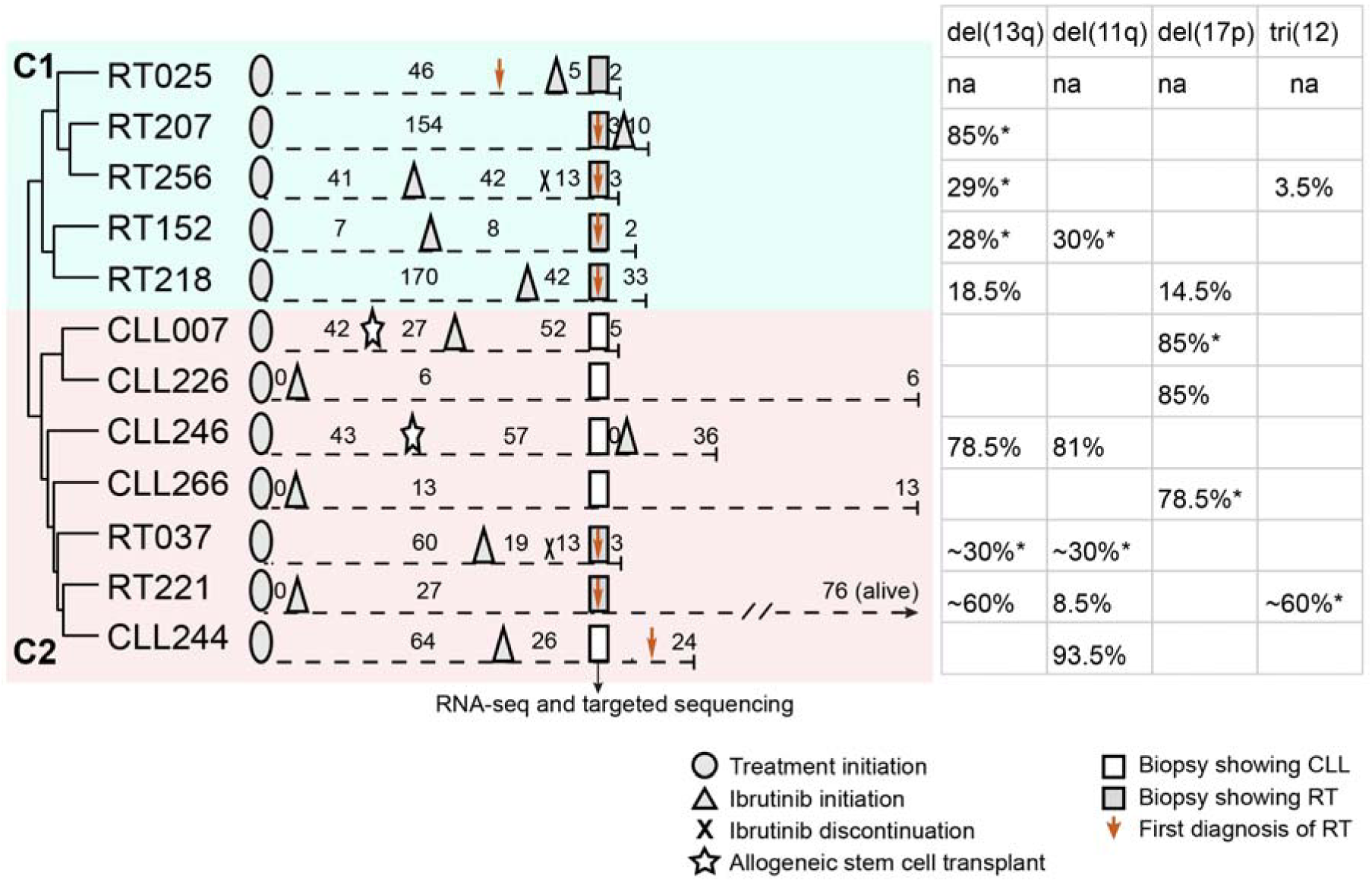
A schematic representation of treatment history and FISH-identified chromosome abnormalities for the 12 patients. The status of being RT vs. CLL is based on biopsy-proven-diagnosis and reviewed by an expert hematopathologist. Seven had confirmed RT (large cell lymphoma histology) and the other five had CLL. Of the five CLL cases, CLL244 developed RT 14 months after tissue biopsy showed CLL. The number indicates time in months, not drawn at the same scale between samples. Biopsy used for RNA-seq and targeted sequencing is marked. Additional Information about treatment history was provided in Table S1. FISH results from bone marrow, lymph node or peripheral blood are available for 11 patients (except RT025), showing the % of nuclei with the indicated abnormality; asterisk indicates abnormalities that are also identified by PatternCNV from targeted sequencing data. The two clusters (C1 and C2) were based on unsupervised clustering of RNA-seq, which used log_2_(counts-per-million) from 12,935 expressed genes on autosomes. del(13q), deletions of 13q; del(11q), deletions of 11q; del(17p), deletions of 17p; tri(12), trisomy 12.

### 2.2. Targeted DNA sequencing and data preprocessing

The Tempus xO oncology exome sequencing panel interrogates genetic alterations in 1,711 cancer-related genes (17). Lymph node formalin-fixed paraffin-embedded (FFPE) samples were used. Targeted DNA was sequenced to 126 base pairs (bp) in paired-end mode on the Illumina HiSeq platform. In addition, five of the 12 patients (RT025, RT218, CLL007, CLL226, and CLL244) had peripheral blood samples available. For these five patients, genomic DNA was sequenced with the same panel, which was used to filter out variants and copy number variation of potential germline origin for all 12 patients.

Raw reads from targeted sequencing were aligned to the hg38 genome reference using Novoalign (v3.05.0) (http://www.novocraft.com/products/novoalign/), with the option “-i 180,46 -t 150”. Duplicates were marked using Picard MarkDuplicate command (v2.9.0, https://broadinstitute.github.io/picard/).

### 2.3. Variant calling and CNA detection

Point mutations and small INsertions and DELetions (INDELs) were identified with Genome Analysis Toolkit (GATK) MuTect2 (v3.7) at default parameter setting (18). Raw variants were annotated for variant quality using GATK walker VariantAnnotator, followed by functional annotation with the Biological Reference Repository (BioR, v4.1.2) (19). BioR integrates variant information from 1000 Genomes Project (20), ExAC (21), gnomAD (22), ClinVar (23) and Human Gene Mutation Database (HGMD) (24), and uses Clinical Annotation of Variants (CAVA, v1.2.0) (25) to stratify variants into categories based on the predicted severity of impact. Only mutations with median or high impact were retained.

Annotated raw variants were subjected to a series of quality filtering. The criteria include minimal allelic (≥10) and overall depth of coverage (≥30), average read mapping quality (≥30), proximity to homopolymer run (≤5), number of reads with MAPQ=0 (≤10), and strand bias (FS≤60). In addition, calls from potential paralogs were removed using R package mapexr (26), as well as those from low-complexity regions (https://github.com/lh3/varcmp/raw/master/scripts/LCR-hs38.bed.gz) (27) and SNP clusters (≥3 SNPs within 20-bp window).

Common variants were eliminated if they had >0.05% minor allele frequencies (MAF) in any of the following germline variant databases: 1000 Genomes Project, ExAC, ESP, and gnomAD. Finally, the retained variants were flagged as being potential germline if they were also identified in the normal. The genes carrying candidate somatic variants were compared to a list of 74 recurrently mutated genes previously identified in CLL (28-31). Only those in the list were further considered.

CNAs in patient tumors were identified with PatternCNV (v1.1.2) (32), which estimates region-level log ratios over the average coverage profiles of normal samples. CN loss was inferred if log_2_(ratio) < -1.0 and CN gain if log_2_(ratio) > 0.585 (https://cnvkit.readthedocs.io/en/stable/calling.html).

### 2.4. RNA sequencing (RNA-seq) and data analysis

Whole-exome capture RNA-seq library preparation and sequencing were performed by Tempus AI, Inc. (Chicago, IL) (33). The RNA samples were then hybridized to the xGen Exome Research Panel (Integrated DNA Technologies, Coralville, IA) and libraries were sequenced in paired-end mode on the Illumina HiSeq 4000 System.

RNA-seq data were analyzed with MAP-Rseq pipeline (v3.1.3) (34). In brief, paired-end reads were aligned to hg38 genome reference using STAR (v2.5.2b) (35). After excluding those with mapping quality below 20, gene raw counts were estimated using featureCounts in the Subread package (v1.6.5) (36), based on the Ensembl gene annotation v78. Gene expression level was quantified as reads per kilobase of exon per million mapped reads (RPKM), using total reads mapped to protein-coding genes on autosomes and chrX.

We next examined the heterogeneity of gene expression across the 12 CLL/RT cases. Protein-coding genes with no mapped reads in all the samples were excluded, and raw counts of the remaining genes were normalized with the Trimmed Mean of M-values (TMM) method in edgeR (v3.16.5) (37). A total of 12,935 expressed genes were selected from autosomes that had raw counts ≥10 and RPKM ≥1 in at least two samples. The counts-per-million (CPM) value from TMM normalization was log_2_ transformed and then used in unsupervised clustering with the pheatmap R package (v1.0.8). The analysis identified two main clusters, C1 and C2, and two subgroups within C1 (C1a and C1b), with distinct expression profiles. Principal component (PC) analysis of the same 12,935 genes was performed using the “prcomp” function in R.

Between the clusters, differentially expressed (DE) genes were identified using edgeR, at the cutoff of FDR ≤0.05 and fold-change ≥2. Subsets of the most discriminatory genes were extracted based on k-means clustering of DE genes with pheatmap. The subsets represented genes up-regulated in cluster C1, in the 3-sample subgroup C1a within C1, or in C2. For the discriminatory genes, enriched pathways were identified using the Enrichr package (https://maayanlab.cloud/Enrichr/) (38).

Deconvolution of bulk RNA-seq data was performed with CibersortX (39). LM22 leukocyte gene signature matrix representing 22 immune cell types was downloaded from CibersortX website (https://cibersortx.stanford.edu). Deconvolution was performed using a customized signature matrix. To generate the signature matrix, expression data from the two B-cell types (naive and memory B cells) within LM22 were replaced with microarray expression data from tumor B-cell in CLL (GSE21029) (40), as described in (41), and from DLBCL (GSE12195) (42). The microarray data was normalized using Affymetrix’s MAS 5.0 expression measure (mas5) in the affy R package. Gene expression was quantified as transcripts-per-million (TPM) using edgeR. Deconvolution was performed using the customized signature matrix, with batch correction mode on (“B-mode”) and quantile normalization disabled. For a given cell type, the difference in abundance between C1 and C2 was tested using Wilcoxon signed-rank test (two.sided, p value <0.1).

## 3. Results and Discussion

### 3.1. Marked heterogeneity of gene expression across CLL/RT

The clinical and therapeutic history for the 12 CLL/RT cases is illustrated in Figure 1. Additional information is provided in Table S1. To identify gene expression changes among RT and CLL, we analyzed RNA-seq data generated from nodal tissue. Unsupervised clustering of 12,935 expressed protein-coding genes from autosomes revealed two major clusters (C1 and C2) with distinct expression profiles (Figure S1A). Cluster C1 includes five of the seven RT. There are 2 subclusters within C1, where the 3-sample subcluster C1a (RT025, RT207 and RT256) showed elevated expression compared to the 2-sample subcluster C1b (RT152 and RT218) for a subset of genes. Cluster C2 includes the other two RT and all five CLL cases. To rule out the possibility that the clustering is due to the chromosomal abnormalities detected by FISH, we excluded 1,734 genes from 13q, 11q, 17p and chromosome 12. Unsupervised clustering of the remaining genes revealed the same pattern with two major clusters and two subcluster within C1. From the 7 RT samples, the analysis identified 3 groups, C1a, C1b and a group of two samples in C2, indicating a high level of heterogeneity in gene expression among RT. Using unsupervised clustering of RNA-seq data, a recent study classified 36 RT into 5 groups, with 3 to 13 patients with RT per group (4).

Within C2, CLL244 developed RT 14 months after tissue biopsy showed CLL (Figure 1). Interestingly, CLL244 had an expression profile more similar to the two RT cases, rather than to the other CLL within C2 (samples within circle, Figure S1B). The results suggest a possibility that RNA-seq can potentially identify expression signature for RT prior to the diagnosis, thus complementing the pathological evidence to better characterize CLL/RT. A previous study indicates that subclones having already acquired expression features of RT can still be under CLL phase for years before transformation occurs (7).

### 3.2. Cluster C1 is preferentially associated with up-regulated genes enriched in key pathways

To identify cluster and subcluster specific expression signature, we performed DE analysis, followed by k-means clustering of the DE genes. The analysis identified 1,431 discriminatory genes from autosomes, including 840 genes up-regulated in the 3-sample subcluster C1a, 474 up-regulated in C1 and 117 up-regulated in C2 (Figure 2A). Thus, 92% (1,314) of the genes showed increased expression in C1, all being RT, particularly within C1a. A previous study based on bulk RNA-seq also revealed a clear trend of more genes being upregulated in RT compared to CLL (7).

**Figure 2.**
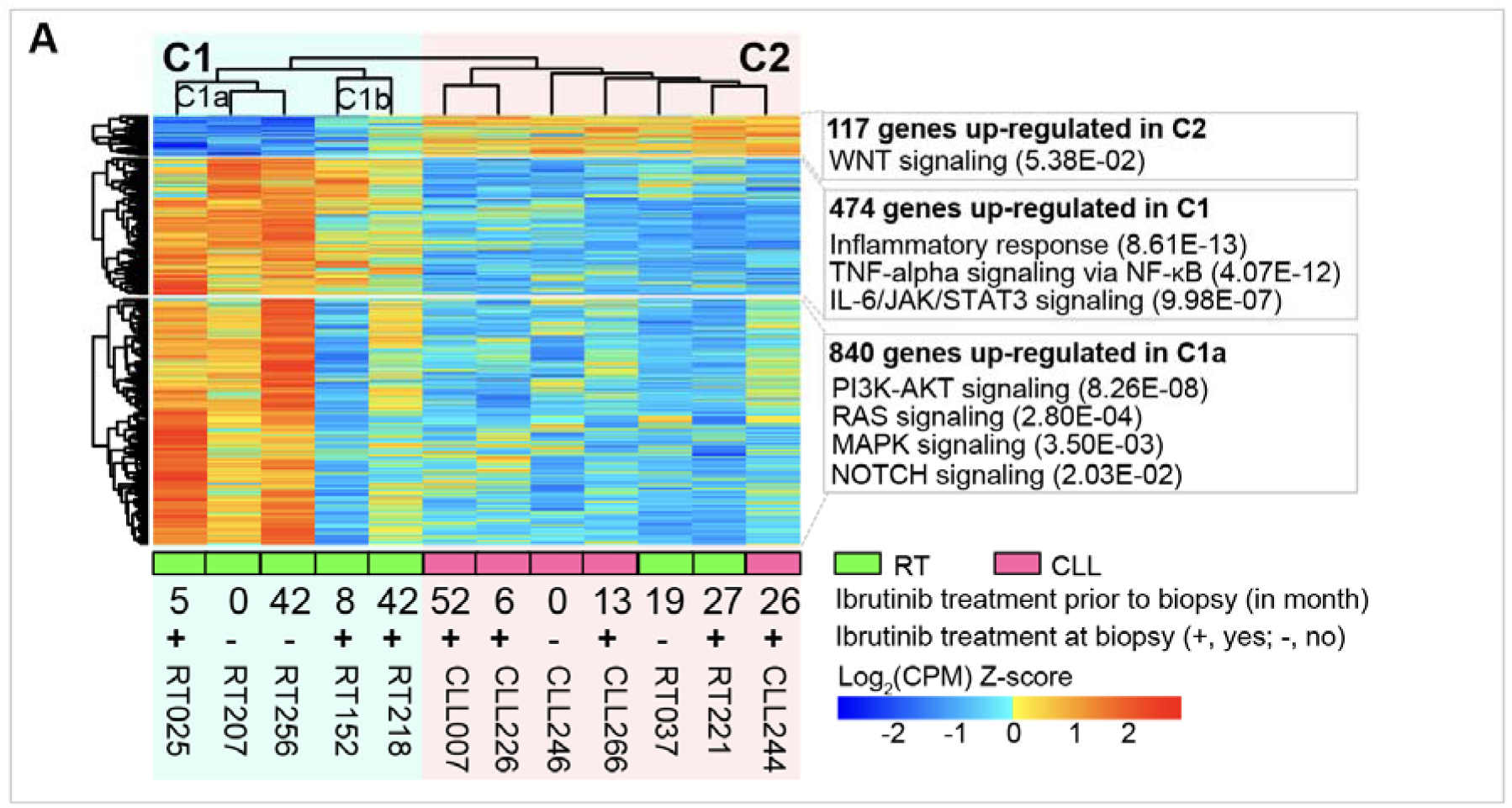
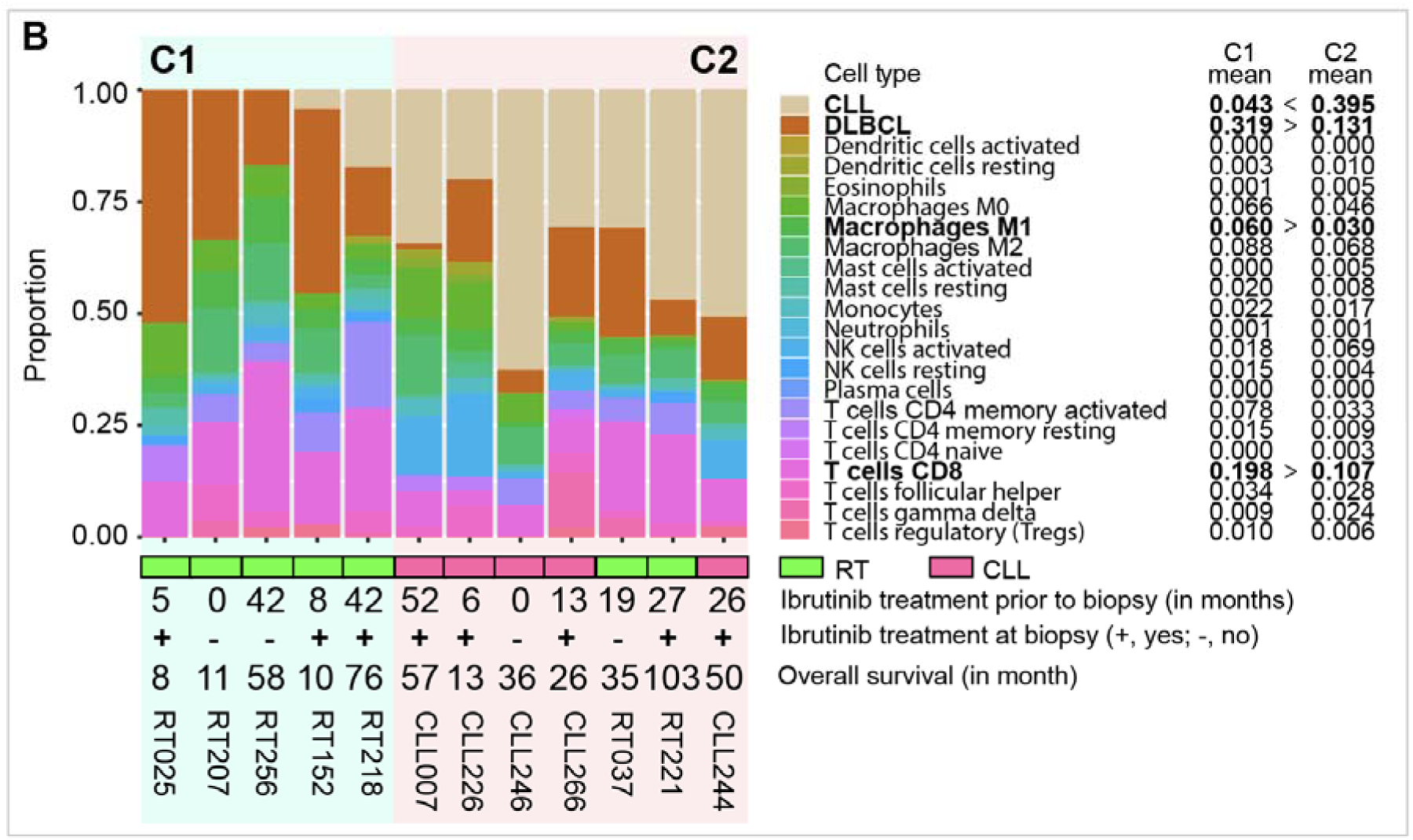
Distinct cellular composition and expression profile of C1 vs. C2. (**A**) Heatmap showing expression changes of DE genes between C1 and C2. The 1,431 discriminatory genes were selected based on k-means clustering of DE genes. Of these, 117 genes were down-regulated in cluster C1, 474 were up-regulated in C1, and the other 840 were up-regulated in the 3-sample subgroup C1a. Enrichr was used to identify the enriched pathways for the three gene lists. (**B**) Proportion of predicted cell types based on CIBERSORTx deconvolution of bulk RNA-seq data. Prior to CIBERSORTx analysis, the two B-cell types (naive and memory B cells) in Leukocyte gene signature matrix (LM22, https://cibersortx.stanford.edu) were replaced with tumor B cells from CLL (GSE21029) and DLBCL patients (GSE12195). The four cell types with significant differences in abundance between C1 and C2 are in bold (p 0.1, Wilcoxon signed-rank test).

These DE genes are enriched in key pathways (Table S2, Figure 2A). Many of them were previously known to play important roles in CLL pathogenesis or transformation (3, 4, 7, 9). Specifically, genes up-regulated in C2 are enriched in WNT signaling, one of the key pathways known to be dysregulated in CLL (43, 44). In this pathway, *ROR1*, *WNT3* and *LEF1* are the three core genes with known roles in CLL (44); all three were up-regulated in C2 relative to C1 (fold change: 2.6 – 12.7; FDR: 6.07E-10 – 1.69E-02). On the other hand, genes up-regulated in C1 are mostly enriched in immune response related pathways, such as TNF-alpha signaling (45, 46), IL-6/JAK/STAT3 signaling (47) and apoptosis (48). Finally, genes up-regulated in C1a are enriched in pathways involved in RT, such as PI3K-AKT signaling (49) and NOTCH signaling (50), or in pathways associated with aggressive clinical course in CLL, such as RAS signaling (51) and MAPK signaling (8, 51). The results suggest that there is marked heterogeneity in gene expression among the RT and CLL patients and that the genes with altered expression are enriched in key pathways involved in CLL or RT.

### 3.3. Major differences in cellular composition between C1 and C2

To understand whether the bulk RNA-seq based clustering of C1 and C2 reflects the potential differences in cellular composition, we performed deconvolution using the customed gene signature matrix. The bulk RNA-seq was deconvoluted into 20 types of non-B cells, as well as tumor B cells from CLL and DLBCL. Clustering based on the predicted cell proportions revealed the same two clusters. Between C1 (RT only) and C2 (a mixture of RT and CLL), there were major differences in the abundances of tumor B cells from CLL vs. DLBCL (Figure 2B). Notably, within C1, the 3-sample subcluster C1a that had the most up-regulated genes had no detectable tumor B cells from CLL, compared to the other two RT in C1b having 4% and 17% tumor B cells from CLL. However, in C2, the two RT who clustered together with CLL had much higher proportions of tumor B cells from CLL (31% and 47%).

In addition, we observed higher abundances of macrophages (M1) and CD8+ T cells (∼2-fold enrichment, p <0.1) in C1 (all RT). Across the 12 samples, RT had higher proportions of macrophages M1 (1.5-fold) and CD8+ T cells (2.8-fold) compared to CLL (Figure 2B). Using immunohistochemistry staining, we previously found that RT cases had higher levels of CD163+ macrophages compared to CLL (2). A recent study using single-cell RNA-seq of longitudinal samples also revealed higher proportions of T cells and macrophages but a lower proportion of B cells in RT compared to the CLL phase (52). Thus, our results support the earlier findings of diversified immune cell composition between RT and CLL (2, 52), with higher levels of macrophages and CD8+ T cells in RT.

Finally, we compared the overall survival of patients in C1 vs. C2 (Table S1, Figure 2B). Not surprisingly, patients in C1 had a shorter OS than those in C2 (median: 11 vs. 36 months), supporting the poor prognosis reported for RT. Also, CLL patients in C2 tend to have an aggressive clinical course, with three of the five cases having an OS of 13-36 months. We found that OS was negatively correlated with the estimated abundance of tumor B cells from DLBCL in the 12 RT/CLL (Spearman R=-0.80, p= 0.0028, Figure 3), and separately in C1 (R=-1, p= 0.017) and C2 (R= -0.68, p= 0.11). Indeed, the three RT in C1 that had the highest abundance of DLBCL cells (>30%) had the shortest OS (8-11 months).

**Figure 3.**
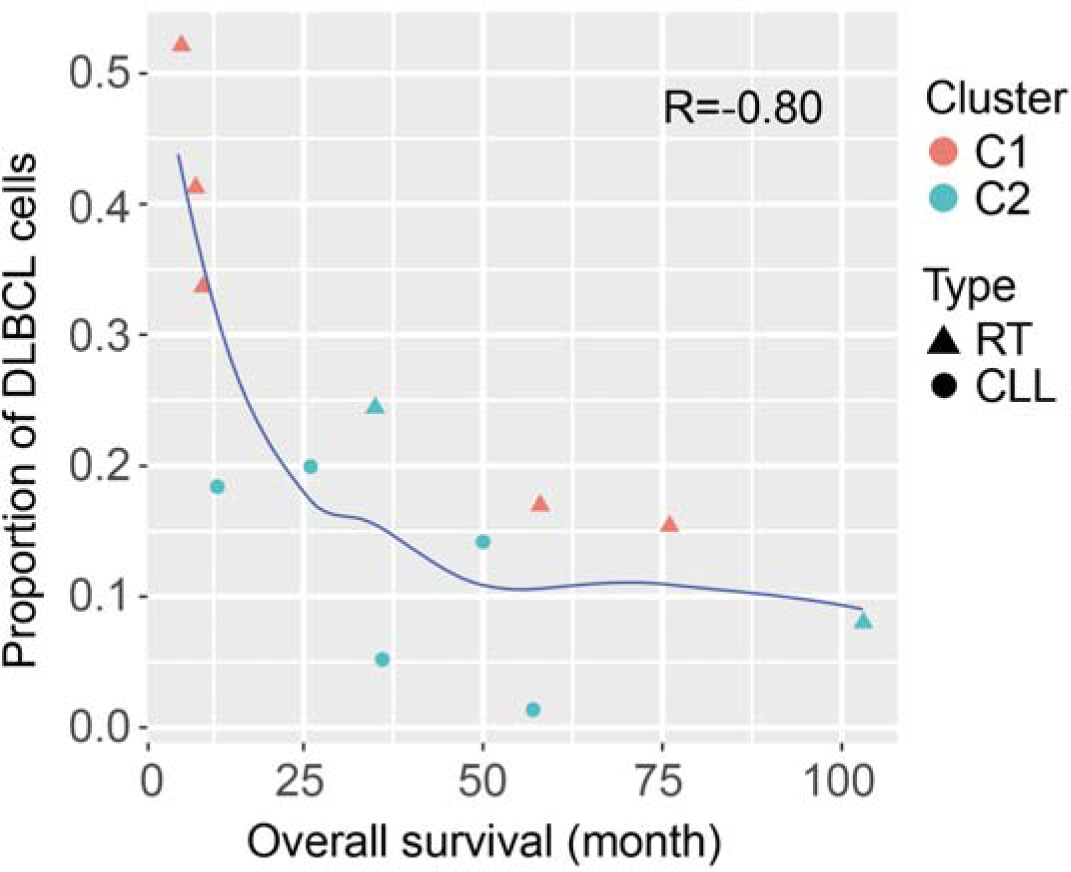
Scatter plot of overall survival vs. abundance of DLBCL cells. Abundance of DLBCL cells in bulk RNA-seq was estimated with CIBERSORTx. Overall survival represents the time from initiation of ibrutinib treatment to death or the last follow-up. Solid line denotes loess curve. Overall survival was inversely correlated with the abundance of DLBCL cells (Spearman R=-0.80, p= 0.0028).

### 3.4. Recurrent CNAs largely occurred in cluster C2

Fluorescence in situ hybridization (FISH) was performed on 11 of the 12 cases (Table S1). Using bone marrow, lymph node or peripheral blood (Table S1), FISH revealed recurrent chromosomal abnormalities commonly observed in CLL (29, 53-56), including deletions of 13q (7 cases), deletions of 11q (5 cases), deletions of 17p (4 cases), and trisomy 12 (2 cases) (Figure 1). Since FISH is a targeted assay, we extended our analyses to identify CNAs from the targeted sequencing data. We confirmed the FISH-identified CNAs in seven cases (Figure 1).

To understand whether there are recurrent CNAs that preferentially occurred in either of the clusters, we extracted C1-enriched CNAs, i.e., those with a prevalence >20% higher in C1 than in C2, and vice versa. The analysis identified 9 C1- and 42 C2-enriched CNAs that were each linked to the respective gene harboring the CNA. We filtered out *CDK11A* gene that showed both CN gain (in 3 cases) and loss (in 2 cases). All the remaining 50 genes showed CN gains (Figure 4A). Five of the genes (n=9) carrying CN gain enriched in C1 are from 1q21q22. Of the other genes (n=41) with CN gain enriched in C2, 73% (n=30) are from 17q12q25, followed by 9q34 (n=5) and 22q13 (n=3). Plot of gene expression levels revealed a tendency of CN gains associated with increased gene expression (Figure 4B).

**Figure 4.**
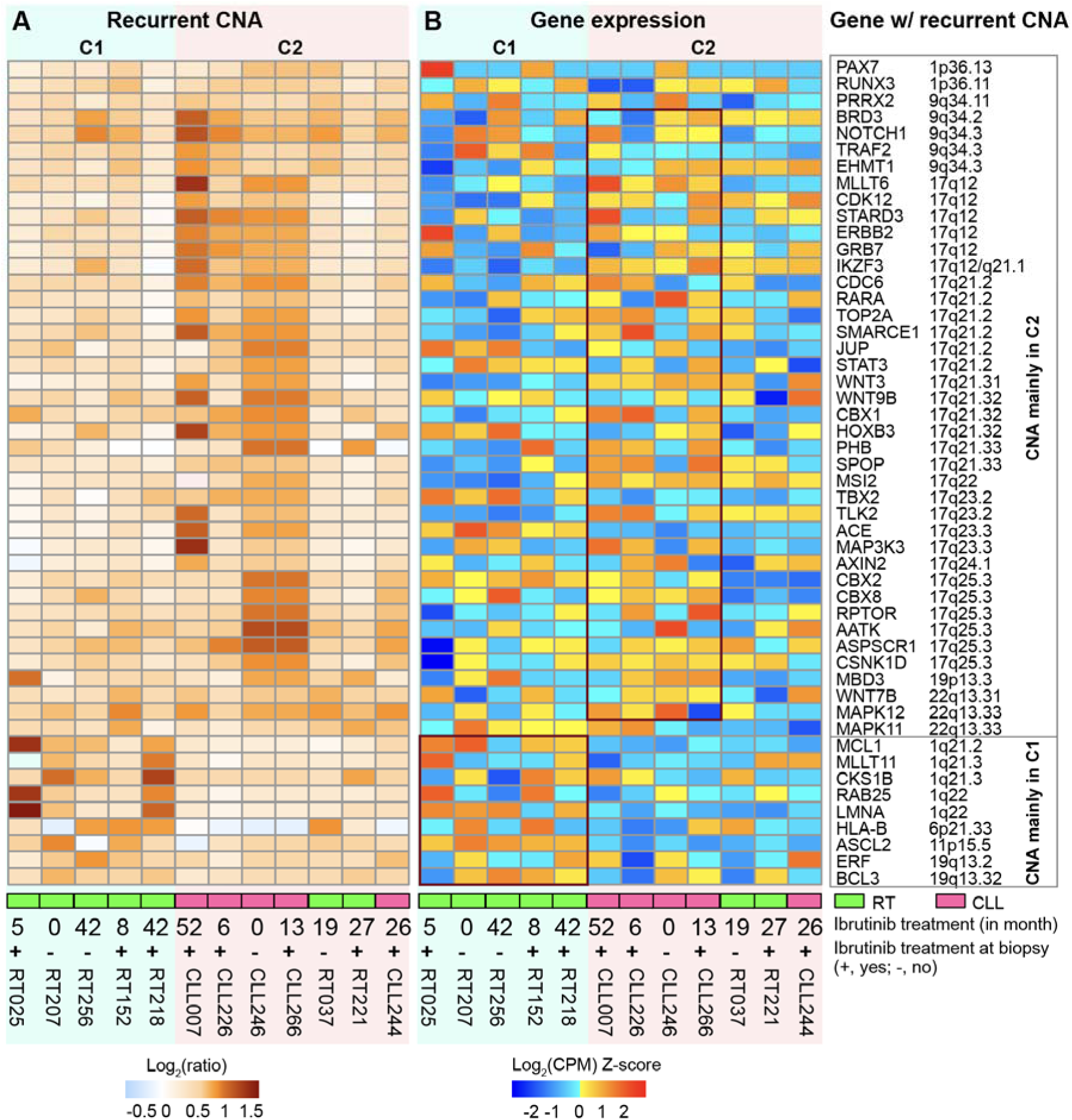
Recurrent CNAs and expression changes in target genes. (**A**) CNAs enriched in cluster C1 or C2. (**B**) Heatmap showing gene expression changes across samples. CNAs were identified with PatternCNV. CN loss was inferred if log_2_(ratio) -1.0 and CN gain if log_2_(ratio) 0.585. CNAs enriched in C1 are defined as those with a prevalence >20% higher in C1 than in C2. The same criteria were used to extract CNAs enriched in C2. At this cutoff, there are 9 and 41 genes that carried CNAs preferentially occurred in C1 and C2, respectively. The genes and their chromosome locations are showed at the right side. There is a trend of association between CN gains and increased gene expression (boxed in B).

Within the above four regions carrying C1- or C2-enriched CNAs, the Tempus xO oncology panel covered a total of 122 genes (Table S3). We further checked the CN status over all the 122 genes. Overall, 53-71% of the genes harbored recurrent CNAs present in at least two cases, the majority of which were enriched, as defined above, or exclusively present in one of the two clusters. Specifically, 14 genes on 1q21q22 carried recurrent CNAs, 11 of which were enriched (n=5) or only present (n=6) in C1. On the other hand, on 17q12q25 two-thirds (30+16+6) of the genes had recurrent CNAs (all CN gains), predominantly being enriched (n=30) or only present (n=16) in C2. C2 includes two RT and five CLL. However, the two RT each had only two genes with C2-enriched CNAs, indicating that CN gains on 17q12q25 preferentially occurred in CLL, not in RT (Figure 4A).

In CLL or RT, CNAs were reported previously in similar regions (Table S3). Using whole-exome sequencing (WES) (57, 58) and whole-genome sequencing (WGS) from CLL (57), two studies have identified small CNAs, mostly with low frequencies of 1-2%. These focal CNAs were 14.7-187 kb in size, which are much smaller than the CNAs we identified. A rare (<2%) 17q arm-level gain was also discovered in CLL (57). In addition, using Affymetrix SNP array (9, 10), two other studies identified CNAs, almost exclusively in RT cases (Table S3). The CNA on 17q previously reported in RT (9) appears to be part of the CNA highly enriched in CLL cases within C2. In summary, we identified recurrent CNAs in four regions, largely 17q gain that preferentially occurred in CLL within C2.

### 3.5. Putative somatic mutations in driver genes

To understand whether there are genes that are preferentially mutated in either of the two clusters, we identified somatic mutations in driver genes. Of the 15 genes with putative somatic mutations, four (*CHD2*, *CNOT3*, *NOTCH1* and *TLK2*) each had mutation in a single sample, and the other 11 each had mutations in 2 to 5 samples (Figure S2). Among the latter, *TP53* had mutations in majority of the cases, mainly missense mutations as previously reported (8, 59). *IKZF3* and *BCOR*, both encoding transcription factors involved in hematologic malignancies (60, 61), harbored mutations in cluster C2. On the other hand, mutations in *CARD11* and *CREBBP* were identified in cluster C1 (all RT) alone. Notably, a previous study with longitudinal samples identified *CREBBP* mutations only at RT phase or during disease progression (7). Given the small sample size, further study with large cohort is needed to validate whether these genes are mutated more frequently in RT or CLL.

## 4. Conclusions

This study revealed a noticeable heterogeneity in gene expression and genetic lesion between RT and CLL, albeit in a small cohort. We observed two major clusters, which also showed differences in cell composition. The vast majority of the DE genes showed elevated expression in RT-only cluster C1, more pronounced for the 3-sample subcluster. Genes up-regulated in this subcluster are enriched in PI3K-AKT, NOTCH, and RAS/MAPK signaling pathways. In addition, there is a strong tendency of recurrent CNVs occurring in C2, mainly gains of 17q12q25 in the CLL cases. Patients in C1 had a shorter OS compared to C2, and OS showed negative correlation with the abundance of DLBCL tumor B cells. Further work is needed to validate the findings in large cohort.

## Supplementary Materials

Figure S1: Gene expression variation in CLL/RT. (A) Unsupervised clustering of expressed genes and (B) Principal component (PC) analysis; Figure S2: Genes recurrently mutated in CLL/RT; Table S1: Clinical information and treatment history for the 12 patients; Table S2: Enriched pathways associated with discriminatory genes; Table S3: Summary of CNAs in the four regions.

## Author Contributions

Conceptualization, H.Y. and W.D.; Investigation, C.Z-M., L.B.B., and D.L.V.D.; Resources, S.A.P., E.W.K., Y.W., M.S., R.H., S.J.K., P.J.H., N.E.K., E.B., and S.L.S.; Data analysis, S.T., H.W., Y.L., H.J-L., E.J., and F.L.; Writing – Original Draft Preparation, S.T., H.W., H.Y. and W.D.; Writing – Review & Editing, all authors. All authors have read and approved the manuscript.

## Institutional Review Board Statement

The genetic testing was conducted according to the guidelines of the Declaration of Helsinki and approved by the Institutional Review Board (or Ethics Committee) of Mayo Clinic (protocol: IRB 18-001468).

## Informed Consent Statement

The patient consent form was waived due to the retrospective nature of this genomics study.

## Data Availability Statement

Data presented in this study can be requested from the corresponding authors.

