## Supplemental table and figure for "Genomic profiling reveals molecular heterogeneity in patients with Richter transformation (RT) and chronic lymphocytic leukemia (CLL)"

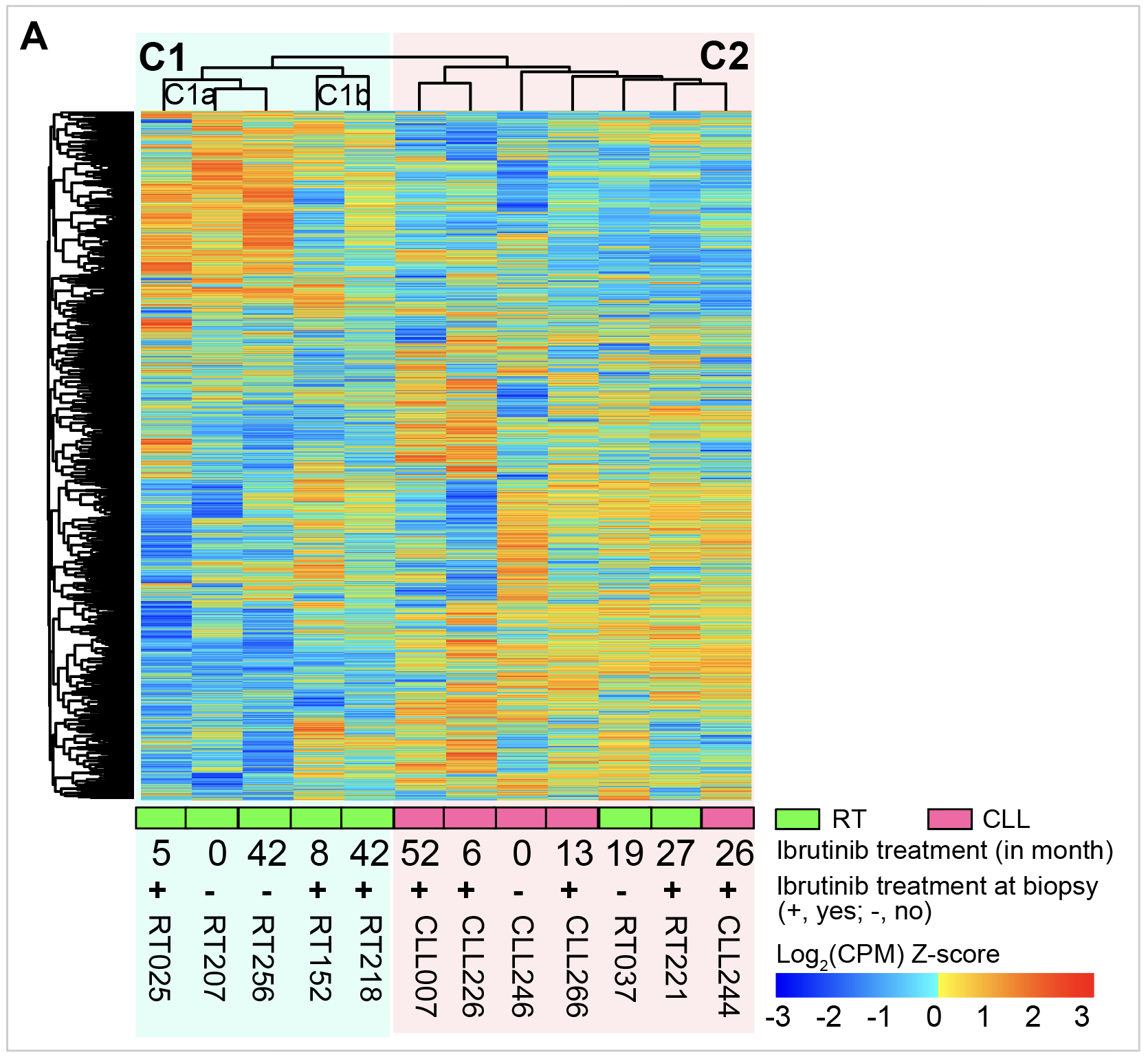

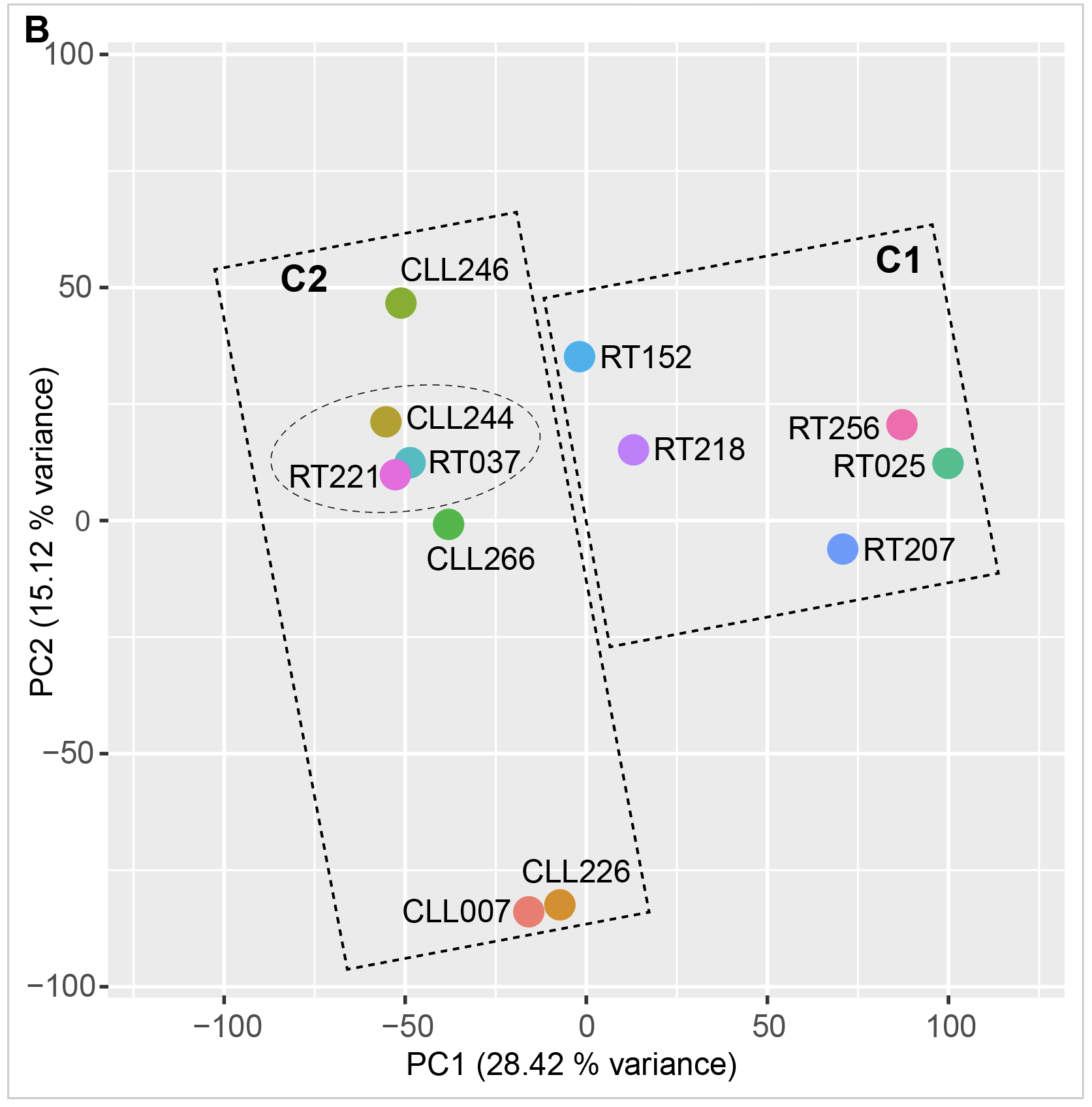

**Figure S1**. Gene expression variation in CLL/RT. (**A**) Unsupervised clustering of expressed genes. A total of 12,935 protein-coding genes from autosomes were used. They had RPKM $\geq$1 and raw counts $\geq$10 in at least two samples. Unsupervised clustering was performed with the R package pheatmap, using log_2_(counts-per-million) after TMM normalization. The 12 CLL/RT samples were grouped into two major clusters, C1 and C2. Within C1, there is a 3-sample subcluster (C1a) that showed elevated expression compared to the 2-sample subcluster (C1b) for a subset of genes. (**B**) Principal component (PC) analysis. PC analysis was performed using the “prcomp” function in R, taking the same 12,935 expressed genes. Within cluster C2, CLL244 developed RT 14 months after tissue biopsy showed CLL. This sample had expression profiles more similar to the two RT (in circle), rather than to the other CLL. PC1 and PC2, principal components 1 and 2.

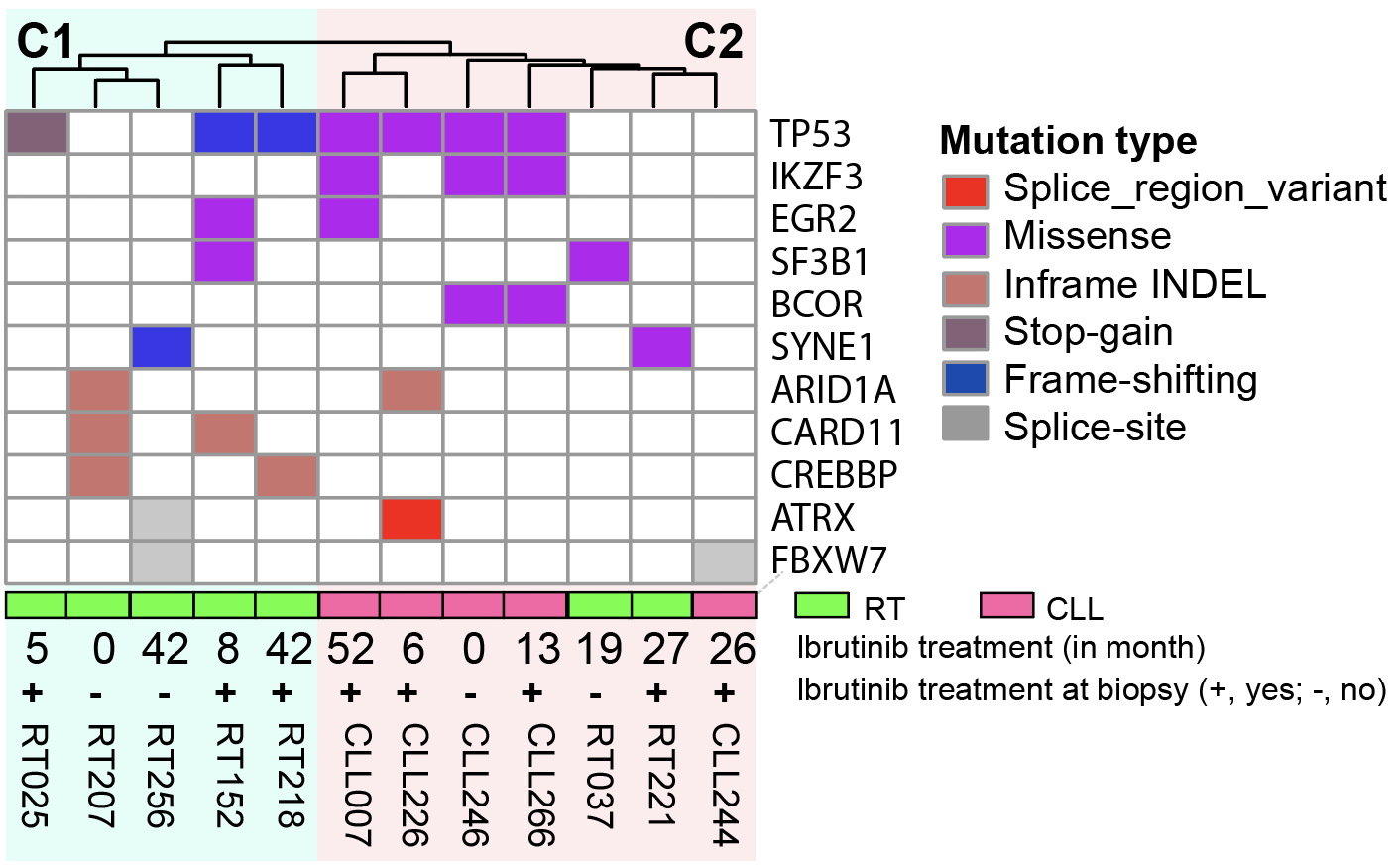

**Figure S2**. Genes recurrently mutated in CLL/RT**.** Raw variants were identified using GATK MuTect2 from targeted sequencing data and stratified into categories based on the predicted impact from CAVA annotation. Raw variants were quality filtered and common variants were eliminated if they had >0.05% minor allele frequency (MAF) in any of the germline variant databases (1000 Genomes Project, ExAC, ESP, and gnomAD), as well as variants that are also present in the normal controls. Shown are the 11 driver genes that had mutations in $\geq$ 2 samples. See Figure 1 legend about sample and treatment information.

**Table S1.** Clinical information and treatment history for the 12 patients

| Sample ID | Sex | TP53 mutation | IGHV mutation | FISH results | Treatment history | OS (month) | Comments about CLL |
| --- | --- | --- | --- | --- | --- | --- | --- |
| RT025 | M | NA | mutated | NA | FCR -> R-CHOP -> radiation-> R-ICE salvage-> pembrolizumab- >ibrutinib added to pembrolizumab (ibrutinib started in 04/2017) | 8 |  |
| RT207 | M | unmutated | NA | del(13q) (bone marrow) | PCR->PAR -> ibrutinib -> R-CHOP | 11 |  |
| RT256 | M | unmutated | NA | del(13q) (peripheral blood and bone marrow); tri(12) (peripheral blood) | R-CVP->R-CP-> Ofatumumab-> Ibrutinib (09/2013 through 03/2017)->Medrol+Rituximab; Ofatumumab | 58 |  |
| RT152 | M | mutated | unmutated | del(13q) and del(11q) (peripheral blood and bone marrow) | Chlorambucil/Gazyva-> Ibrutinib (ibrutinib since 04/05/2017) | 10 |  |
| RT218 | M | NA | unmutated | del(13q) and del(17p) (bone marrow) | FC->PCR->Cytoxan plus rituximab->Revlimid/dexamethasone->FCR->ofatumumab->R/Medrol->ibrutinib (started in 12/2013, continued all the way before biopsy) | 76 |  |
| CLL007 | M | NA | NA | del(17p) (lymph node) | PC->Cyclophosphamide, mitoxantrone, vincristine, and prednisone, maintenance rituximab->Bendamustine->FCR-lite->Radiation therapy->Allogeneic stem cell transplant (2012) ->Ibrutinib (stared in 04/2014, stopped 02/2015)->R-ESHAP->radiation to mass->Ibrutinib (restarted in 12/2015)->Pembrolizumab->Ibrutinib, Obinutuzumab->Venetoclax in combination with ibrutinib->Obinutuzumab added to ibrutinib, venetoclax and prednisone | 57 | relapsed CLL |
| CLL226 | M | unmutated | unmutated | del(17p) (abdominal mass) | ibrutinib (started in 10/2017, on hold since 3/16/18)->Rituximab and Solu-Medrol | 13 | less response on ibrutinib, with concern for progression |
| CLL246 | F | NA | NA | del(13q) and del(11q) (peripheral blood) | FCR->Rituxan/solumedrol->Rituximab / bendamustine->Alemtuzumab, rituximab, Solu-Medrol->Bone Marrow Transplant – Allogeneic stem cell transplant (2013)-> ibrutinib | 36 | purine analog-refractory CLL |
| CLL266 | F | mutated | unmutated | del(17p) (bone marrow) | Ibrutinib (6/30/2017-7/2018) | 26 | relapsed/refractory CLL |
| RT037 | M | unmutated | mutated | del(13q) and del(11q) (peripheral blood and bone marrow) | Leukeran->rituximab->bendamustine/rituximab->ibrutinib (started in 06/2015, then stopped ibrutinib and switched to idelalisib in 01/2017)-> idelalisib with rituximab for 10 months->venetoclax for 3 months | 35 |  |
| RT221 | M | unmutated | unmutated | del(13q), del(11q), and trisomy 12 (peripheral blood) | ibrutinib (started in 12/2015) and rituximab->stopped ibrutinib for a few days in 12/2017, and then restarted around 12/2017->stopped ibrutinib 3 days prior to biopsy | 103 |  |
| CLL244 | M | unmutated | unmutated | del(11q) (peripheral blood and bone marrow) | FCR->Bendamustine/Rituximab->Ibrutinib (started in 2/2016, stopped in 12/2017)->Obinutuzumab/Methylprednisolone, ibrutinib restarted (01/2018) | 50 | relapsed CLL |

Samples are listed in the same order as shown in Figure 1. Overall survival (OS) was defined as the time from initiation of ibrutinib treatment to death or the last follow-up. NA, data not available

**Table S2**. Enriched pathways associated with discriminatory genes

| Term | Adjusted p-value | Source | No. gene |
| --- | --- | --- | --- |
| PI3K-AKT signaling pathway | 8.26E-08 | KEGG | 840 |
| Rap1 signaling pathway | 8.26E-08 | KEGG | 840 |
| Ras signaling pathway | 2.80E-04 | KEGG | 840 |
| MAPK signaling pathway | 3.50E-03 | KEGG | 840 |
| NOTCH signaling pathway | 2.03E-02 | KEGG | 840 |
| cAMP signaling pathway | 2.73E-02 | KEGG | 840 |
| Inflammatory response | 8.61E-13 | MSigDB | 474 |
| TNF-alpha signaling via NF-κB | 4.07E-12 | MSigDB | 474 |
| IL-6/JAK/STAT3 signaling | 9.98E-07 | MSigDB | 474 |
| Apoptosis | 8.44E-06 | MSigDB | 474 |
| Allograft rejection | 3.11E-03 | MSigDB | 474 |
| Hematopoietic cell lineage | 3.85E-03 | KEGG | 474 |
| IL-17 signaling pathway | 1.04E-02 | KEGG | 474 |
| P53 pathway | 2.05E-02 | MSigDB | 474 |
| Wnt signaling pathway | 5.38E-02 | KEGG | 117 |

Pathway analysis was performed using Enrichr (https://maayanlab.cloud/Enrichr/). Of the three gene lists, 840 genes are up-regulated in the 3-sample subcluster C1a, 474 genes are up-regulated in C1, and 117 genes are up-regulated in C2. See Figure 2A about samples in each cluster. KEGG, Kyoto Encyclopedia of Genes and Genomes.

**Table S3**. Summary of CNAs in the four regions

| Cytoband | 1q21q22 | 9q34 | 17q12q25 | 22q13 |
| --- | --- | --- | --- | --- |
| Total no. gene | 24 | 15 | 76 | 7 |
| Genes with CNA | 22 | 13 | 59 | 5 |
| Shared | 3 | 3 | 6 | 2 |
| C1 only | 8+6 | 0 | 0 | 0 |
| C1 enriched | 5 | 0 | 0 | 0 |
| C2 only | 0 | 5+0 | 7+16 | 0 |
| C2 enriched | 0 | 5 | 30 | 3 |
| Region with CNA (hg38) | chr1:146,156,130-156,572,614 | chr9:127,786,079-137,834,963 | chr17:37,687,321-82,832,484 | chr22:45,344,545-50,270,337 |
| Focal recurrent CNA | 33 kb (146,156,130-146,189,400); 5.56 Mb (150,575,185-156,139,153) | 1.04 Mb (128,683,847-129,722,362); 4.07 Mb (133,764,025-137,834,963) | 11.94 Mb (37,687,321-49,622,886); 1.48 Mb (57,256,643-58,734,232); 4.16 Mb (61,400,045-65,558,646); 7.51 Mb (75,320,357-82,832,484) | 632 kb (45,344,545-45,976,844); 310 kb (49,960,871-50,270,337) |
| CLL WES (n=934) and WGS (n=163) (Knisbacher, et al. 2022) | 1q22; CN gain (14.7 kb, <2% CLL) | 9q34.3; CN loss (67.2 kb, ~2% CLL) | 17q21.32; CN gain (115.3 kb, ~2% CLL); also 17q arm-level gain (1.6% CLL) | 22q13.2; CN gain (54.4 kb, 5-6% CLL) |
| CLL WES (n=392) (Mosquera Orgueira, et al. 2019) |  | 9q34.4; CN gain (187 or 84.7 kb, 1.5-2% CLL) | 17q25.3; CN gain (40.1 kb, 1.5% CLL) | 22q13.33; CN gain (40.6 kb; 1% CLL) |
| Affymetrix SNP Array in CLL (n=9) and RT (n=51) (Fabbri, et al. 2013) | 1q21.1q42.12; CN gain (2 RT) | 9q13q34.3; CN gain (1 RT); CN loss (1 CLL) | 17q21.32q25.3; CN gain (3 RT) | 22q13.32; CN loss (1 RT) |
| Affymetrix SNP Array in CLL (n=343) and RT (n=59) (Chigrinova, et al. 2013) | 1q21.1q32.1; CN gain (15% RT) | 9q33.2q34.3; CN loss (15% RT) |  |  |

Three regions had CNAs enriched in cluster C2 (mostly CLL) and one had CNAs enriched in C1 (all RT). Total no. gene, number of genes covered by the Tempus xO oncology panel. Genes with CNAs were broken into “Shared”, “C1 only”, “C1 enriched”, “C2 only” and “C2 enriched”. Shared, a CNA is present in both C1 and C2 but not enriched in either; C1 only, CNA is present in 1 (the first number) or 2 (the second number) cases from C1 but in none from C2; C1 enriched, CNA is present in $\geq$3 of the 12 cases and over-represented in C1 (prevalence >20% higher than in C2; see Figure 4 legend). The same criteria apply to the extraction of C2 only and C2 enriched CNAs. Four previous studies reported CNAs in similar regions.
